# Alcohol, smoking, and brain structure: common or substance specific associations

**DOI:** 10.1101/2024.09.25.24313371

**Authors:** Vera Thornton, Yoonhoo Chang, Ariya Chaloemtoem, Andrey P. Anokhin, Janine Bijsterbosch, Randi Foraker, Dana B. Hancock, Eric O. Johnson, Julie D. White, Sarah M. Hartz, Laura J. Bierut

**Affiliations:** Department of Psychiatry, Washington University School of Medicine, St. Louis, Missouri, USA; Department of Radiology, Washington University School of Medicine, St. Louis, Missouri, USA; Department of Medicine, Washington University School of Medicine, St. Louis, Missouri, USA; GenOmics and Translational Research Center, RTI International, Research Triangle Park, North Carolina, USA; Fellow Program, RTI International, Research Triangle Park, North Carolina, USA

## Abstract

Alcohol use and smoking are common substance-use behaviors with well-established negative health effects, including decreased brain health. We examined whether alcohol use and smoking were associated with the same neuroimaging-derived brain measures. We further explored whether the effects of alcohol use and smoking on the brain were additive or interactive.

We leveraged a cohort of 36,309 participants with neuroimaging data from the UK Biobank. We used linear regression to determine the association between 354 neuroimaging-derived brain measures and alcohol use defined as drinks per week, pack years of smoking, and drinks per week × pack years smoking interaction. To assess whether the brain associations with alcohol are broadly similar or different from the associations with smoking, we calculated the correlation between z-scores of association for drinks per week and pack years smoking.

Results indicated overall moderate positive correlation in the associations across measures representing brain structure, magnetic susceptibility, and white matter tract microstructure, indicating greater similarity than difference in the brain measures associated with alcohol use and smoking. The only evidence of an interaction between drinks per week and pack years smoking was seen in measures representing magnetic susceptibility in subcortical structures. The effects of alcohol use and smoking on brain health appeared to be additive rather than multiplicative for all other brain measures studied. 97% (224/230) of associations with alcohol and 100% (167/167) of the associations with smoking that surpassed a p value threshold are in a direction that can be interpreted to reflect reduced brain health. Our results underscore the similarity of the adverse associations between use of these substances and neuroimaging derived brain measures.

## Introduction

Alcohol consumption and cigarette smoking are common substance-use behaviors, and both are associated with numerous adverse health consequences including heart disease, lung illnesses, cancer, and increased mortality [1–6]. Alcohol consumption and smoking have also been identified as risk factors for dementia [7].

Consistent with the increased risk for dementia, alcohol consumption and cigarette smoking are associated with differences seen across multimodal neuroimaging. Alcohol consumption is associated with global and regional decreases in brain volume [8–10]. Similarly, cigarette smoking is associated with decreased white and gray matter volume as well as numerous regional differences [11,12]. Overall, these decreases in brain volume are often considered adverse consequences of alcohol consumption and cigarette smoking [13–16]. Our goal is to build on this work by exploring whether the neural correlates associated with alcohol use and smoking are largely similar or different across a large set of neuroimaging measures. In addition to global measures of brain volume, there are regional cortical measures of volume, thickness, and area, as well as subcortical volume derived from T1 MRI and subcortical measures of magnetic susceptibility derived from T2* MRI. Measures of white matter integrity are derived from diffusion weighted MRI (dMRI) and resting functional connectivity derived from resting-state functional MRI (rfMRI).

In addition, although alcohol use and smoking often co-occur [17], prior studies of associations of these substances with neural imaging characteristics controlled for co-use, but did not specifically test whether there was an interaction between these substances. An important question is whether the addition of smoking, given the same level of alcohol use, influences the relationship between alcohol and global and regional brain differences, and vice versa for smoking.

The UK Biobank (UKB) enables a well-powered investigation of the neural correlates of alcohol and smoking behaviors through its unprecedented scale, with neuroimaging currently available for tens of thousands of participants. Beyond the neuroimaging data, the UKB includes rich demographic questionnaires, lifestyle features including alcohol and smoking behaviors, genetic characterizations, and patient records through the National Health Service [18]. We analyzed UKB data to take a data-driven approach to examine the associations between alcohol use and smoking behaviors across a comprehensive set of neuroimaging measures to explore the following questions: 1) are the same or different brain regions associated with alcohol use and smoking? and 2) is there evidence of an interaction between alcohol use and smoking with respect to the associations seen with neuroimaging phenotypes?

## Materials and Methods

### Participants

Between 2006 and 2010, over 500,000 participants ages 40 to 69 enrolled in the UKB at 22 centers across the United Kingdom. Participants attended a baseline appointment and responded to a detailed survey on demographics and lifestyle [18].

Since 2014, a subset of participants returned for neuroimaging [19]. The National Health Service North West Centre for Research Ethics Committee granted ethical approval for the UKB (Ref: 11/NW/0382). All participants provided informed consent per UKB procedures. Analyses were conducted under UKB Resource Application Number 48123.

We included all UKB participants with imaging data available as of Spring 2023.

Participants of all races and ethnicities were included. We excluded participants who withdrew consent or had a neurological condition that could affect brain structure. We examined relatedness, and in cases where relationships were 3^rd^ degree or closer, we randomly selected one of the individuals to include for analysis. Participants who were missing information on sex, age, head size, imaging site, imaging date, or rfMRI motion were excluded. We further excluded participants who did not currently consume alcohol but who formerly drank. This resulted in a final cohort of 36,309 participants. See Supplement for a further description of participant inclusion/exclusion rules per the Strengthening the Reporting of Observational Studies in Epidemiology (STROBE) guideline [20]. The final count of participants by MRI sequence is given in the Supplement.

### MRI data acquisition and processing

UKB participants were imaged at one of four sites with identical equipment and following standardized procedures. The MRI data were passed through a quality control and analysis pipeline to tabulate summary measures of brain structure and function called imaging-derived phenotypes (IDPs) [21,22].

### Selection of IDPs for analysis

We selected a subset of IDPs that capture a range of brain measures while avoiding redundant measures. Specifically, our analysis included 354 IDPs: 4 represent total brain volumes from T1 MRI; 186 represent grey matter volume, area, and thickness in 62 cortical regions derived from T1 MRI using the Freesurfer Desikan Killiany parcellation [23]; 36 represent regional subcortical volumes derived from T1 MRI using the Freesurfer ASEG parcellation [23,24]; and 14 IDPs are derived from T2* MRI and represent magnetic susceptibility in subcortical brain structures. We also included 108 IDPs derived from diffusion MRI representing four measures reflecting structural integrity in 27 white matter tracts. These IDPs are the diffusion tensor imaging (DTI) outputs fractional anisotropy (FA) and mean diffusivity (MD), and neurite orientation dispersion and density (NODDI) generated the measures intracellular volume fraction (ICVF) and isotropic volume fraction (ISOVF). We adopted a previous data-reduction approach of the rfMRI IDPs based on independent component analysis (ICA) to obtain six independent components (ICs) representing broad patterns of connectivity [25]. See Supplement for additional information.

### Measures of alcohol use and cigarette smoking

Participants reported their drinking frequency via a survey at the baseline and subsequent imaging visits. Participants were asked to estimate how many drinks they consumed in a typical week (for those who drank on a daily to weekly basis) or in a typical month (for those who drank monthly or less) in standard units of red wine, white wine, fortified wine, beer and cider, spirits, or other (such as alcopops). If alcohol intake varied, participants were instructed to consider their typical week or month within the past year. To derive a standardized measure of drinks per week, we summed alcohol consumption across different drink types to get total units of drinks consumed per week or month. For those who provided a monthly estimate, we converted the monthly totals to weekly estimates by dividing the amount by 4.3. Those who reported no alcohol consumption in the past 12 months were queried to determine if they never drank or formerly drank and now stopped. Those who never drank were assigned 0 drinks per week, and those who formerly drank were dropped from the analysis because of the known misclassification of their alcohol consumption. Although we acknowledge that drinking behavior can vary over the life course, we used this measure of past 12 month alcohol use as a proxy for lifetime alcohol consumption [26]. See Supplement for additional details on how we defined the drinks per week variable.

Participants who endorsed current or former daily or near daily smoking were queried about age of smoking onset and recency and quantity of cigarettes smoked per day. We used these data to calculate “pack years,” a measure of lifetime exposure to smoking. Those who never smoked or smoked fewer than 100 cigarettes in their lifetime were categorized as never smoking and assigned 0 pack years. We assigned 1 pack year to those who endorsed occasional smoking with more than 100 cigarettes over their lifetime, but never smoked daily. See Supplement for further details on how the pack years variable was derived.

### Additional covariates for analysis

The UKB contains a wealth of measures that could be included in our model. To select covariates for the model, we referenced UKB neuroimaging literature [27], which recommended the inclusion of sex, age, head size, imaging site, imaging date, and rfMRI derived motion. We further included income, educational attainment [28–30] and health-related variables of body mass index (BMI), history of diabetes, and systolic and diastolic blood pressure (BP) [31,32] as these have been shown to be associated with neuroimaging characteristics [31–33]. Because we included all participants regardless of race or ethnicity, we included the first 10 genetic principal components.

### Missing survey data

Measures provided during the imaging appointment were used as the variables for analysis. Data missing at the imaging visit were backfilled with the value from the baseline visit. This missing data procedure was applied to drinks per week, pack years, income, educational attainment, BMI, history of diabetes, and systolic and diastolic BP. We performed multiple imputation by chained equations using the classification and regression trees (CART) method for the remaining missing values using the mice package in R [34]. See Supplement for further description of data processing.

### Statistical analyses

Analyses were performed using R statistical software (https://www.r-project.org/) and all code is available on GitHub (https://github.com/BierutLab/ukb_alcohol_smoking). We developed a linear regression model with IDPs as the outcome, and within a single model, we tested the associations with alcohol use as drinks per week and cigarette smoking as pack years. To determine a potential alcohol use-by-smoking interaction, we included an interaction term between drinks per week and pack years. All IDPs and continuous numeric covariates were z-score normalized.

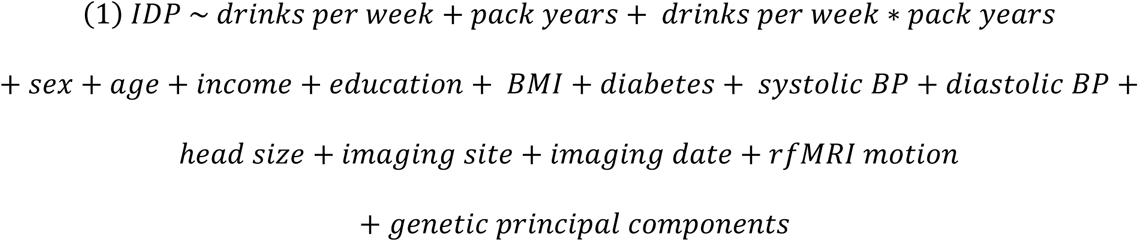

We then calculated a z-score (beta / standard error [se]) for the association of each regional measure with drinks per week and pack years smoking. To quantify the extent to which these associations were similar in direction and magnitude, we determined the correlation between the z-scores of associations with drinks per week and with pack years smoking in sets of IDPs defined by MRI sequence, brain region, and in the case of dMRI, derived measure. We report all results without correction for multiple comparisons.

For analyses with total brain volume, we repeated the analysis without normalizing the brain volumes, drinks per week, or pack years so results can be directly interpreted as volume in mm^3^ per drink per week or per pack year.

## Results

Our final cohort consisted of N=36,309 participants with at least one MRI sequence. Sample demographics are provided in Table 1: 53% of participants were female, 43% were between the ages of 60-69 years at the time of imaging, and 97% reported being “white”, “British”, “Irish”, or “any other white background”. Participants who endorsed current drinking (97% of the cohort) consumed a median of 6 drinks per week. A smaller proportion of the sample (37%) reported current or former smoking; those with a history of daily smoking reported a median of 14.75 pack years.

**Table 1.**
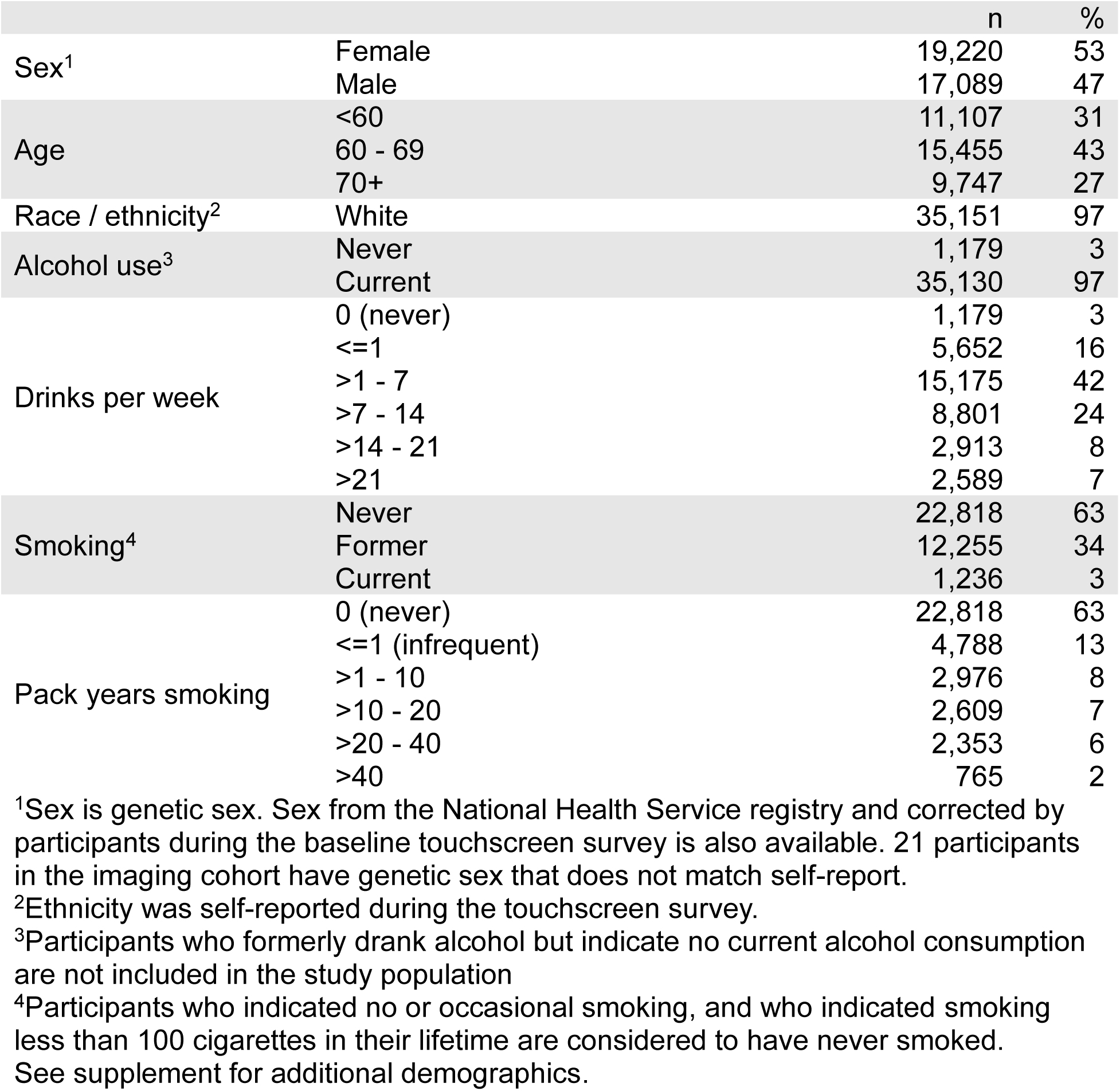
Sample demographics for UK Biobank imaging cohort (N = 36309)

Participants who reported consuming more drinks per week had higher average pack years of smoking. However, the correlation between drinks per week and pack years smoking was low, r = 0.18 (p = 2.8 × 10^-262^).

Drinks per week and pack years were both individually associated within the single model with decreased total brain volume, grey matter volume, and white matter volume. For each additional drink per week, total brain volume decreased by 451 mm^3^ (p=8.14 × 10^-40^). For each additional pack year of smoking, total brain volume decreased by 159 mm^3^ (p=1.53 × 10^-7^) (Table 2). The associations between alcohol and smoking in regional and multimodal IDPs were overwhelmingly in a direction consistent with decreased brain health. Out of 230 IDPs associated with alcohol at the 1% p value threshold level, 224 (97%) were in the direction suggesting decreased brain health while only six were in a direction consistent with improved health. Out of 167 IDPs associated with smoking at the 1% p value threshold level, 167 (100%) were in the direction suggesting decreased brain health.

**Table 2.**
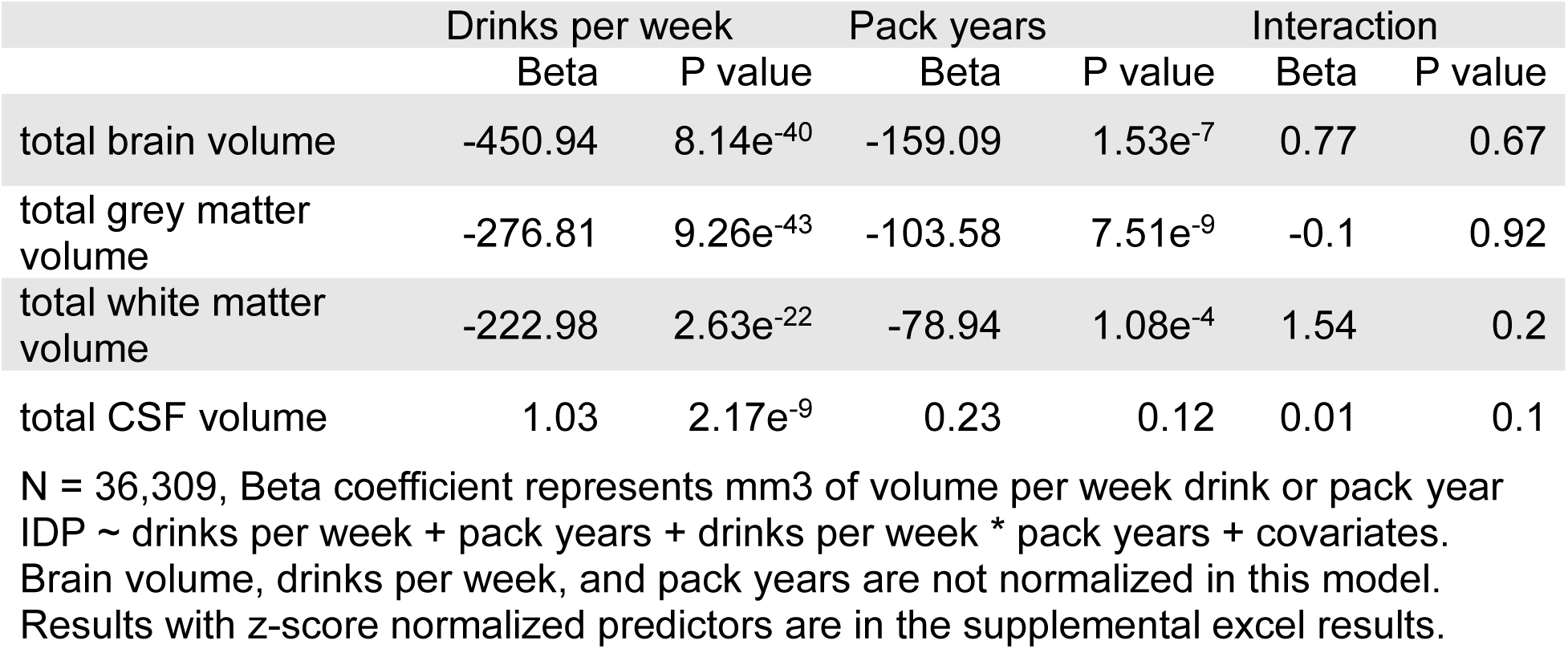
Associations between total brain volumes and alcohol, smoking, and their interaction.

|  | Drinks per week |  | Pack years |  | Interaction |  |
| --- | --- | --- | --- | --- | --- | --- |
|  | Beta | P value | Beta | P value | Beta | P value |
| total brain volume | -450.94 | 8.14e<sup>-40</sup> | -159.09 | 1.53e<sup>-7</sup> | 0.77 | 0.67 |
| total grey matter volume | -276.81 | 9.26e<sup>-43</sup> | -103.58 | 7.51e<sup>-9</sup> | -0.1 | 0.92 |
| total white matter volume | -222.98 | 2.63e<sup>-22</sup> | -78.94 | 1.08e<sup>-4</sup> | 1.54 | 0.2 |
| total CSF volume | 1.03 | 2.17e<sup>-9</sup> | 0.23 | 0.12 | 0.01 | 0.1 |

N = 36,309, Beta coefficient represents mm<sup>3</sup> of volume per week drink or pack year IDP ~ drinks per week + pack years + drinks per week \* pack years + covariates. Brain volume, drinks per week, and pack years are not normalized in this model. Results with z-score normalized predictors are in the supplemental excel results.

To determine whether the associations with alcohol use and smoking were similar or different across a range of brain measures, we calculated a z-score of association of the IDPs with drinks per week and with pack years smoking (Figure 1).

There was high correlation (r = 0.995, p = 0.004) between the z-scores of association with drinks per week and z-scores of association with pack years smoking in the four measures of total brain volume (Figure 1A). In IDPs representing cortical volume, thickness, and area in 62 brain regions defined using the Freesurfer DKT atlas, we found a moderate correlation (r=0.47, p=1.4 × 10^-4^) between the associations of cortical volume and drinks per week and pack years smoking and a moderate correlation (r=0.63, p=4.0 × 10^-8^) for cortical thickness and drinks per week and pack years smoking (Figure 1B). All associations surpassing a p value threshold of 0.01 between alcohol and smoking and IDPs representing cortical volume and thickness have a negative direction of effect consistent with decreased cortical volume and thickness. While we found no correlation (r=-0.02, p=0.88) between the associations between cortical area and drinks per week and pack years smoking, this is likely because the associations with alcohol and smoking both had small effect sizes in these measures. Only 21 surpassed the 1% p value threshold for association with alcohol and four passed for smoking. Of these all had a negative direction of effect.

**Figure 1.**
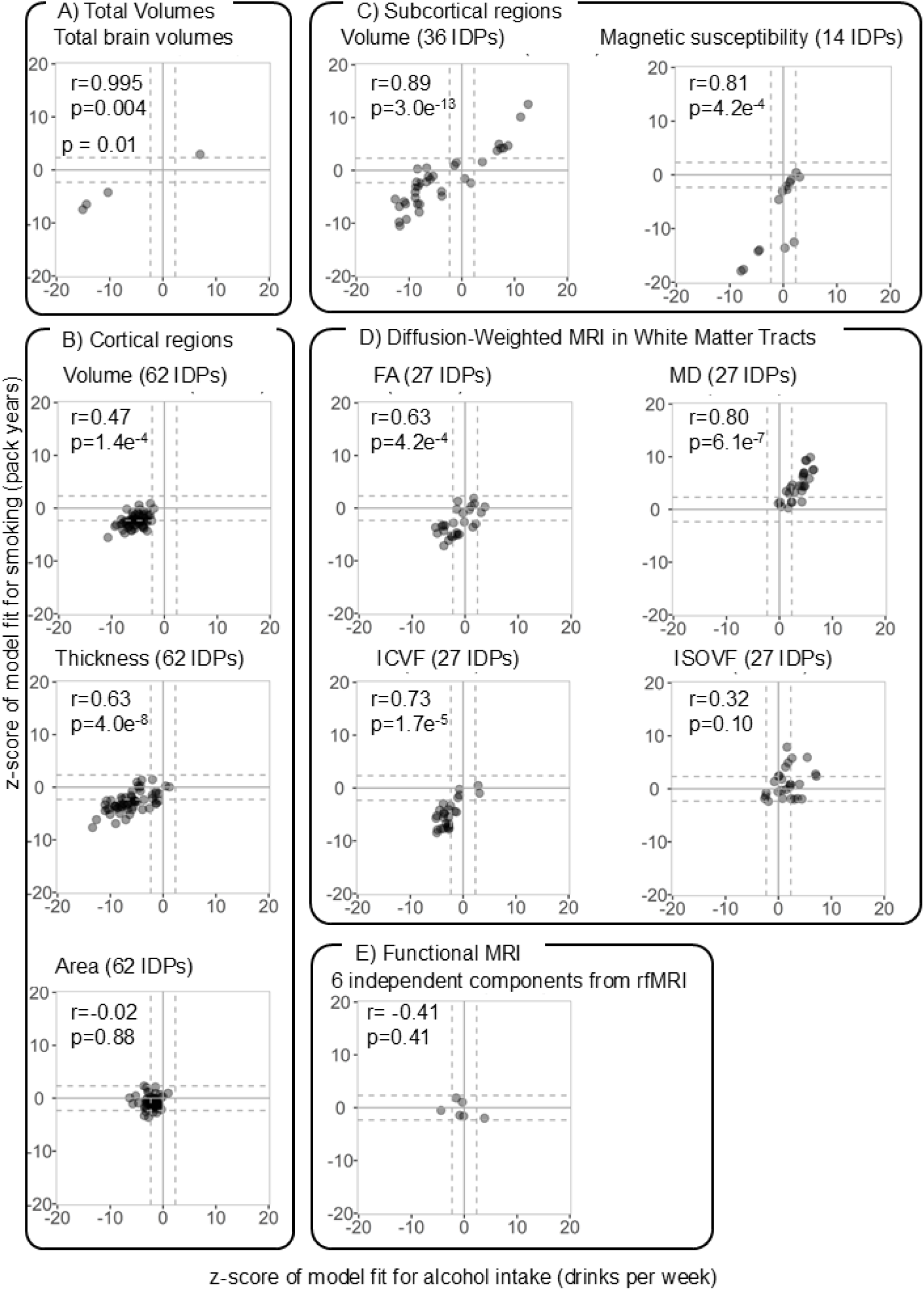
Scatter plots of correlation between z-score of association with alcohol vs smoking for brain measures. Scatter plots to visualize the correlation (Pearson’s product moment correlation, r) between the z-score (beta / se) of association with alcohol (drinks per week) and the z-score of association with smoking (pack years) for each IDP. Dashed grey line represents p = 0.01. Model equation: scaled IDP ∼ scaled drinks per week + scaled pack years + scaled drinks per week * scaled pack years + covariates.

We analyzed 36 IDPs representing volumes of subcortical structures from the Freesurfer ASEG atlas. Correlation between the z-scores of drinks per week and pack years in these structures was high (r = 0.89, p=3.0 × 10^-13^). Direction of effect associated with both drinks per week and pack years was negative (decreased volume) for all IDPs representing tissue volume and was positive for IDPs representing volume of ventricles (Figure 1C). In the 14 IDPs representing magnetic susceptibility in subcortical structures, correlation between the z-scores of drinks per week and pack years was high (r=0.81, p=4.2 × 10^-4^). Four out of the five IDPs surpassing the 1% p value threshold for association with alcohol had a negative direction of effect, and all nine IDPs associated with smoking had a negative direction of effect (Figure 1C). Decrease in these measures is consistent with decreased brain health.

We analyzed 108 IDPs representing four dMRI-derived measures in 27 white matter tracts. In three of the four measures (fractional anisotropy [FA], mean diffusivity [MD], and intracellular volume fraction [ICVF]), correlation between the z-scores of drinks per week and pack years smoking was moderate (FA r = 0.63, p= 4.2 x 10^-4^; MD r= 0.80, p=6.1 x 10^-7^; ICVF r=0.73, p=1.7 x 10^-5^). For both drinks per week and pack years, the direction of effect associated with FA and ICVF was negative and with MD was positive (Figure 1D), which suggests decreased white matter structural integrity across multiple tracts. The ISOVF measures showed less correlation (r=0.32, p=0.1) in the associations with drinks per week and pack years smoking.

We included six independent components (ICs) derived from rfMRI in our analysis. No evidence of correlation was found between z-scores for drinks per week and pack years in these measures (p=0.41) (Figure 1E). Drinks per week was associated with ICs 1 and 3 surpassing p < 0.01, whereas pack years of smoking was not associated with any of the ICs (p > 0.09). We do not interpret the direction of effect associated with ICs in terms of increased or decreased brain health because this is not established. Further results for all IDPs are available in the Supplement.

Our second question was whether there was evidence of an interaction between alcohol use and smoking with respect to the associations seen with the neuroimaging measures. We found no statistical interaction (p > 0.1) between drinks per week and pack years for any of the four measures of total brain volume (Table 2). Out of 354 IDPs, only six had an interaction p value less than 0.01 and only one of these surpassed a Bonferroni threshold of p=4 x 10^-5^ (Figure 2). All six IDPs represented magnetic susceptibility from T2* in subcortical structures (Table 3). Individually, both drinks per week and pack years smoking were associated with hypointensity in T2* measures, suggesting iron deposition and decreased brain health [35–37]. With increasing co-use of alcohol and smoking, the individual contributions of drinks per week and pack years smoking converge. At high pack years smoking, further increasing drinks per week does not contribute to greater hypointensity. Figure 3A shows how the interaction with drinks per week influences the association between pack years and magnetic susceptibility in the left caudate, contrasted to the association between pack years and total brain volume where we find no evidence of an interaction (Figure 3B).

**Figure 2.**
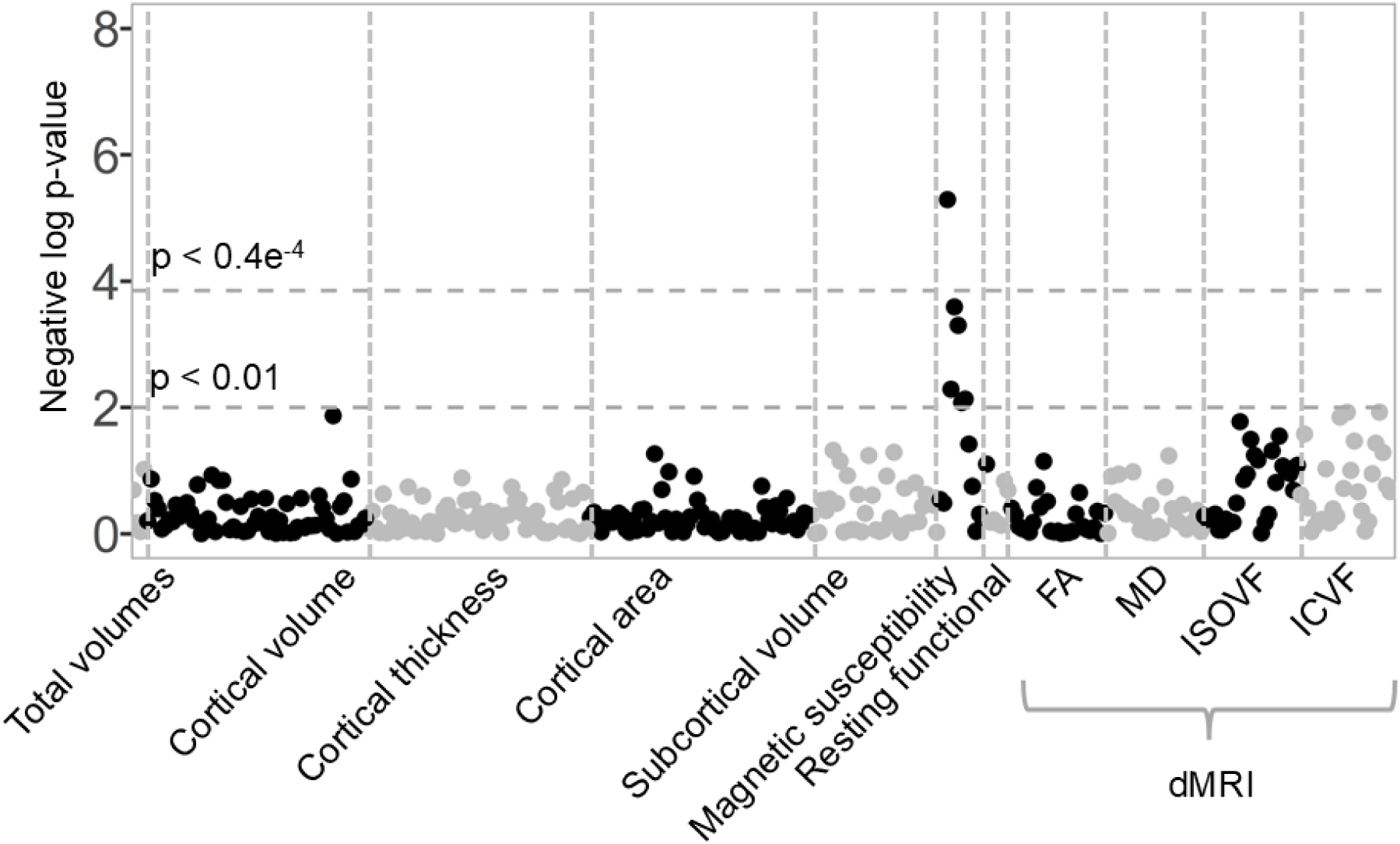
Negative log p value of the interaction term in 354 IDPs. We employed a plot of the negative log of p-values to identify IDPs across all brain measures in which the interaction term reaches any of several levels of statistical significance. We do not find an interaction with p < 0.01 in total brain volume, total grey matter volume, or total white matter volume. IDPs representing magnetic susceptibility from T2* in subcortical structures form a peak.

**Figure 3.**
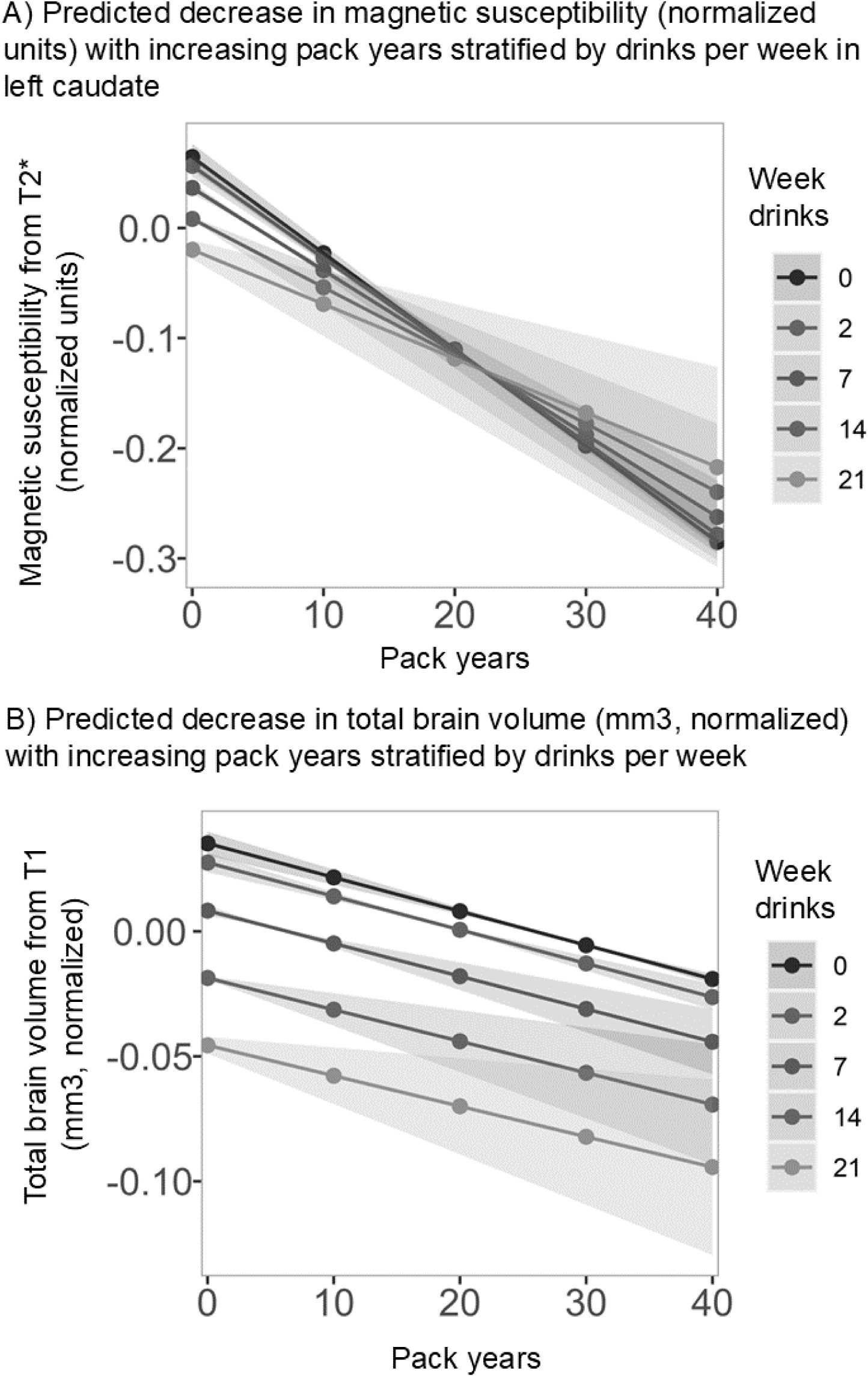
Comparison of the relationship between pack years and brain difference in total brain volume vs. left caudate magnetic susceptibility. A) Plot of total brain volume (mm^3^, normalized) by pack year, stratified by drinks per week. We do not see any evidence of an interaction in which drinks per week affects the association with pack years. B) Plot of magnetic susceptibility in the left caudate (normalized units) by pack year, stratified by drinks per week. In this structure we see evidence of an interaction in which the slope of the relationship with pack years differs depending on drinks per week.

**Table 3.**
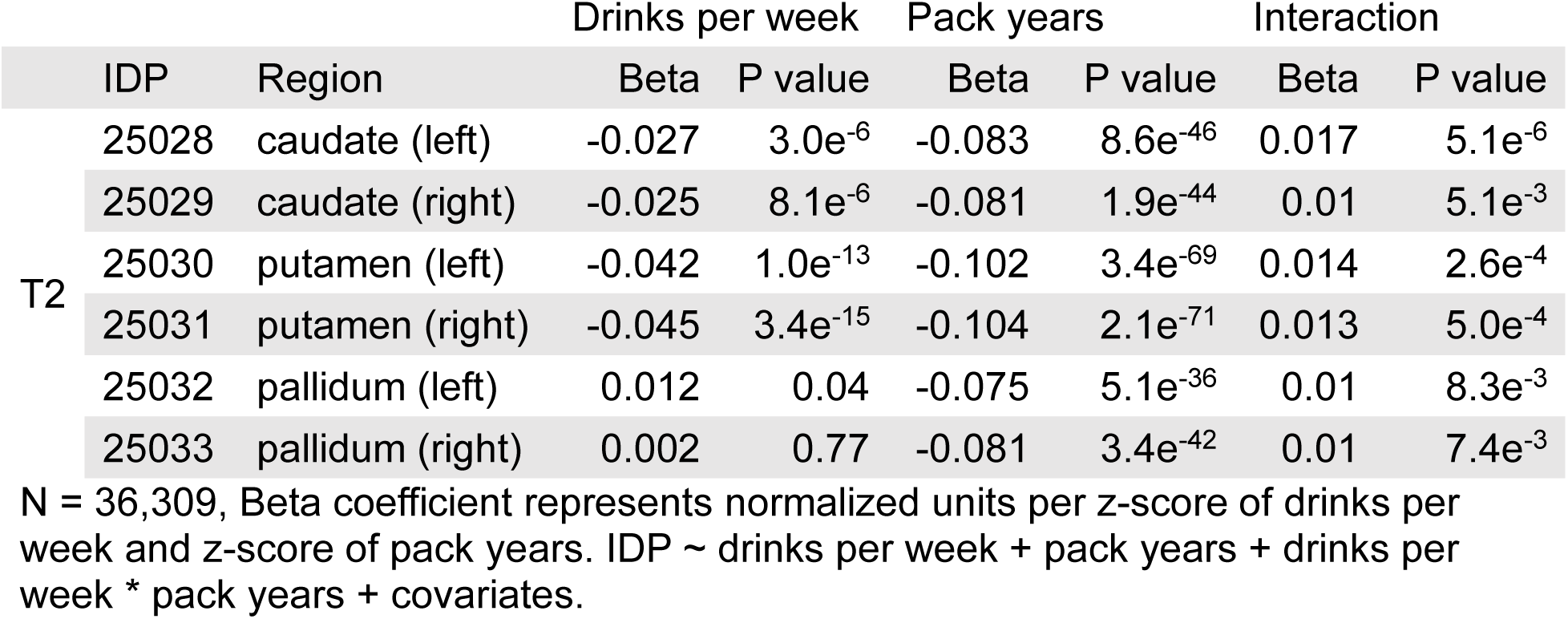
Imaging derived phenotypes (IDPs) with interaction temp p value < 0.01.

## Discussion

Many regions across multiple neuroimaging modalities were associated with both drinks per week and pack years of smoking. The correlation between associations was moderate for cortical volumes. We found high correlation in subcortical volumes, with both drinks per week and pack years associated with decreased tissue volumes. Decreased volume of cortical and subcortical structures along with increased volume of ventricles is plausibly a marker of decreased brain health [38]. Multiple studies have observed decreased total brain volumes associated with both alcohol and smoking. Alcohol and smoking have also been shown to be associated with decreased cortical and subcortical volumes, with these decreases spread diffusely across the brain rather than concentrated in specific regions [8–13]. Our results replicate these findings and indicate broad similarity in the regions that are associated.

In addition, IDPs representing magnetic susceptibility were highly correlated (r = 0.81) with drinks per week and pack years smoking, with all associations reaching the 1% p value threshold level in the negative direction (hypointensity) in the T2* signal.

Hypointensity on T2* suggests iron deposition in brain tissue, which could result from iron storage dysregulation or bleeding into brain tissue such as from micro strokes [35–37]. Associations between alcohol use, hypointensity on T2* in subcortical structures, body iron, and decreased cognitive performance have been reported [16]. We extend this finding in alcohol by assessing the association with smoking as well. We determined that the associations with alcohol consumption measured by drinks per week and cigarette smoking defined by pack years smoked are highly correlated in the subcortex.

dMRI-derived measures were associated with alcohol and smoking in a pattern of decreased fractional anisotropy (FA) and intracellular volume fraction (ICVF) along with increased mean diffusivity (MD), and the correlations in these measures were moderate to high. Decreased FA along with increased MD suggests decreased directionality of water movement through the brain, while decreased ICVF suggests decreased density of neurites [39–41]. The pattern of decreased FA and ICVF and increased MD has been shown to be associated with both alcohol and smoking [8,9,12]. It has also been identified in hypertension, diabetes, and aging, all conditions of decreased brain health [32,39]. Our results replicate these prior findings in alcohol and smoking and the correlations we find support that within each measure, association with alcohol predicts similar association with smoking and vice versa.

Drinks per week was associated with only two out of the six ICs from rfMRI, and we did not find any association between pack years smoking and any of the ICs. It is not surprising then that we found no correlation between the z-scores of alcohol and smoking associations for the rfMRI measures (r = -0.41, p = 0.41).

In addition to exploring the degree to which associations with alcohol and smoking were correlated, we tested for interaction between these behaviors and the neuroimaging measures. We found little evidence of an interaction. Six IDPs surpassed a p value threshold of 0.01 and only one surpassed a Bonferroni correction (0.05/354) for the interaction. All these IDPs represented magnetic susceptibility from T2*. The direction of this effect indicated that those who both drink and smoke heavily will show slightly less deleterious difference than would be expected from adding up the differences associated with each substance individually. It is important not to interpret this as a potential protective effect, because the direct association with each substance is larger in magnitude and in the direction of decreased brain health. Instead, we interpret this finding as a limit on difference in this measure at high consumption of both substances. Given the large population available through the UKB, we interpret our findings to indicate that other than the T2* findings, if any interaction effect exists, it is small.

### Limitations

Despite several strengths, including the large sample size, extensive neuroimaging measures, and joint examination of alcohol and smoking measures, we note some limitations of our study. Alcohol use and smoking are known to be correlated [42] and this introduces a concern for collinearity. We determined that correlation between drinks per week and pack years smoking was minimal in this population (see Supplement). This low correlation assuages concerns that the validity of model results may be impacted by collinearity between the measures of alcohol and smoking.

We have refrained from definitively claiming that regions are not associated with alcohol or smoking because this could be an artifact of our power to detect such an association. Examination of our results shows that in many cases the regions that would reach a given statistical significance threshold for association are physically adjacent to regions that barely fail to meet the threshold. Thus, it is misleading to emphasize the importance and interpretation of a single isolated region that reaches the threshold over one directly adjacent that barely fails. We have reported results with un-adjusted p values and at the 1% threshold to avoid artifacts of this nature. Similarly, this limitation applies to our exploration of the interaction between alcohol and smoking. Although the UKB provides the largest neuroimaging sample to date, we may still be underpowered to detect an interaction effect particularly at the highest levels of alcohol use and smoking, as this represents a relatively small subsample (see Supplement).

Collection of alcohol use and smoking data also has limitations. Participants may incorrectly report their use, leading to mis-classification [43,44]. A lifetime measure of total alcohol consumption, similar to the lifetime measure of pack years smoked, would be ideal to examine associations between brain measures and alcohol, but is not available. Instead UKB participants reported their typical alcohol consumption over the past year. We interpreted this past year measure as being representative of their alcohol consumption across the lifetime, and we acknowledge the potential for misclassification by using this approach [26].

Another important limitation of this work is that results only establish association, they do not indicate causation. Although the brains of participants who consumed alcohol or smoked cigarettes differed, we cannot claim that these differences were caused by alcohol consumption and smoking. An alternative explanation is that these correlates of alcohol use and smoking may represent features that predispose one to use alcohol and smoke [45,46].

Finally, the UKB represents a subset of the UK population willing and able to undergo intensive screening. Participants who returned for a repeat assessment were older, had higher income, higher education, came from areas with lower levels of material deprivation (Townsend deprivation score), and lived closer to the assessment center than those who were invited but did not participate [47,48]. It is also important to note that UKB participants had a much lower prevalence of smoking as compared to the UK population [42,47]. Although we included participants of all races and ethnicities in our study cohort, due to the demographics of the UKB participants, our sample was over 97% white [49].

## Conclusion

The findings of this study support common, rather than distinct, associations of MRI-based brain phenotypes with alcohol and smoking. We found both alcohol and smoking associated with decreased brain volumes and differences suggestive of decreased brain health across the overwhelming majority of MRI derived measures. There was evidence of an interaction in IDPs representing magnetic susceptibility in subcortical structures but little evidence for interaction across all other measures.

## Data Availability Statement

UKB data used in this study are available after application to the UKB.

## Supporting information

Supplemental full results

## Acknowledgements

We would like to thank the research participants who generously shared their health information with the UKB, as well as all administrators and employees of the UKB. We would like to recognize Louis Fox and Sherri Fisher for their support throughout the project.

## Author Contributions

VT conceived the idea, analyzed and interpreted data, prepared figures, and wrote the manuscript. YC and AC contributed to data analysis and interpretation. AC participated in figure preparation. JB and SMH advised and guided data analysis. JB provided curation of UKB imaging data. APA, JB, RF, DBH, EOJ, JDW, and SMH guided the conceptualization and design of the work and interpretation of the data. LJB provided resources, developed and supervised the project, and contributed to and oversaw all conception, design, analysis, interpretation, figures, and writing of the manuscript. All authors reviewed and edited the manuscript.

