## Supplemental methods for "Alcohol, smoking, and brain structure: common or substance specific associations"

Supplemental methods 1. Study inclusion / exclusion

Our cohort includes all participants who completed the first neuroimaging appointment and had data available as of Spring 2023 (N=42,801). Participants of all races and ethnicities are included in the study sample. We kept participants who could be matched to our phenotype data and removed all those who had withdrawn consent. We then excluded participants based on neurological conditions as described in supplemental methods 4 “Exclusion on neurological conditions”. We kept only one person of 3rd degree or closer relative pairs as described in supplemental methods 6 “Exclusion of 3rd degree or closer relatives”. We excluded participants who were missing any of the UKB recommended neuroimaging controls (Alfaro-Almagro et al., 2021) included in our analysis (head size, sex, imaging site, and age). We removed all participants with past alcohol use but who did not currently consume alcohol. The steps of study inclusion / exclusion and number of participants in the cohort at that step are given in supplemental methods 2 “Table of participants in the original UKB imaging sample and through study inclusion / exclusion”.

Supplemental methods 2. Table of participants in the original UKB imaging sample and through study inclusion / exclusion

| Step | N |
|---|---:|
| Imaging cohort | 42,801 |
| Matched phenotype participant ID across imaging and survey | 41,761 |
| Consent withdrawn removed | 41,749 |
| Neurological conditions removed | 40,118 |
| 3rd degree or closer relatives removed | 39,167 |
| Missing UKB recommended neuroimaging covariates removed | 37,512 |
| Participants who formerly drank but do not currently consume alcohol | 36,309 |

Our final cohort is 36,309 participants. We divided the imaging data (IDPs) into sets based on the MRI sequence and processing methods used to derive those IDPs. Within each set, we excluded participants who were missing that IDP sequence. As a result, each sequence-based dataset has a slightly different final number as participants may be missing some sequences and not others. The MRI sequence-based sets and final number of participants in each is given in supplemental methods 3 “Table of final N in MRI sequence defined sets”.

Supplemental methods 3. Table of final N in MRI sequence defined sets

| IDP set | N |
|---|---|
| Total volumes from T1 structural MRI | 35,921 |
| Regional measures from T1 structural MRI | 35,919 |
| T2* magnetic susceptibility | 34,065 |
| Diffusion weighted MRI (dMRI) | 35,835 |
| Resting-state functional MRI (rfMRI) | 33,171 |

Supplemental methods 4. Exclusion on neurological conditions

UKB Data Field 20002 (https://biobank.ndph.ox.ac.uk/showcase/field.cgi?id=20002) provides codes for self-reported non-cancer illnesses. Participants were asked in the baseline survey if a doctor had ever told them they have a serious illness. If they responded yes, they were asked about the illness during an interview with a trained nurse who assigned it a numeric illness code (https://biobank.ndph.ox.ac.uk/showcase/refer.cgi?id=100235). If there was any uncertainty as to the illness, the participant described it to the nurse who used their expertise to code it correctly. Numeric codes given in the UKB Data Field are matched to the names of the illness or condition with data coding 6 (https://biobank.ndph.ox.ac.uk/showcase/coding.cgi?id=6). If the illness could not be determined well enough to code it during the interview the nurse entered a free text description which was later coded by a physician. If the illness could not be determined after this procedure, it was coded as 99999 “unclassifiable”.

We referenced the codes in Data Field 20002 to exclude participants with certain neurological conditions. We based our list of conditions on the exclusion criteria in Gray 2020 (Gray et al., 2020). The condition name, condition code, and number of participants with that condition are reported in supplemental methods 5. The number of participants excluded for neurological conditions is slightly less than the sum of the numbers with neurological conditions as some participants had multiple conditions.

Supplemental methods 5. Neurological conditions for which participants were excluded and the number of participants excluded for each condition

| Condition | Condition code | N participants | % participants* |
|---|---|---|---|
| Brain / intracranial abscess | 1245 | 5 | 0.01 |
| Brain hemorrhage | 1491 | 19 | 0.05 |
| Cerebral aneurysm | 1425 | 10 | 0.02 |
| Cerebral palsy | 1433 | 3 | 0.01 |
| Chronic degenerative neurological | 1258 | 8 | 0.02 |
| Dementia / Alzheimer’s | 1263 | 14 | 0.03 |
| Encephalitis | 1246 | 29 | 0.07 |
| Epilepsy | 1264 | 240 | 0.57 |
| Guillain-Barre syndrome | 1256 | 21 | 0.05 |
| Head injury | 1266 | 150 | 0.36 |
| Meningioma | 1659 | 15 | 0.04 |
| Meningitis | 1247 | 205 | 0.49 |
| Motor neuron disease | 1259 | 10 | 0.02 |
| Multiple sclerosis | 1261 | 134 | 0.32 |
| Nervous system infection | 1244 | 1 | <0.01 |
| Neurological disease / trauma | 1240 | 13 | 0.03 |
| Other demyelinating disease | 1397 | 8 | 0.02 |
| Parkinson’s | 1262 | 77 | 0.18 |
| Spina bifida | 1524 | 9 | 0.02 |
| Stroke | 1583 and 1081 | 458 | 1.10 |
| Subarachnoid hemorrhage | 1086 | 30 | 0.07 |
| Subdural hematoma | 1083 | 9 | 0.02 |
| Transient ischemic attack | 1082 | 296 | 0.71 |

*Percent of participants is calculated from the number of participants in the cohort (N = 41,749) before removing those with neurological conditions

Supplemental methods 6. Exclusion of 3rd degree or closer relatives

Due to geographic and relational factors there are numerous groups of related individuals in the UK Biobank (Bycroft et al., 2018). We identified relatives of 3rd degree or closer using a kinship matrix provided by the UK Biobank. 1st degree relatives have an expected kinship coefficient of 1/4, 2nd degree relatives have an expected kinship coefficient of 1/8, while 3rd degree relatives have a kinship coefficient of 1/16. To ensure all 3rd degree or closer relatives were identified, we filtered for kinship coefficients equal to or greater than 1/20. Within that group of relatives, we identified related pairs with both individuals in the neuroimaging set. Within those pairs with neuroimaging data, we implemented an algorithm based on the one developed by Hanscombe et al. to drop participants in the order of the most highly interconnected until only pairs of related individuals remain (Hanscombe et al., 2019). We then randomly selected one member of each pair to remove.

Supplemental methods 7. dMRI abbreviations and their meanings

| Abbreviation | Full | What is it? | Interpretation |
|---|---|---|---|
| DTI | Diffusion tensor imaging | Measure of the movement of water in MRI | |
| NODDI | Neurite orientation dispersion and density imaging | An expanded set of measures based on a more sophisticated model of water movement | |
| FA | Fractional anisotropy | Directional coherence of water diffusion | Increase in FA is greater directionality of water movement suggests integrity of white matter tracts consistent with better brain health |
| MD | Mean diffusivity | Undirected diffusion | Increase in MD is decreased directionality of water movement which suggests loss of integrity in white matter tracts |
| ICVF | Intracellular volume fraction | Neurite density | More water in cells suggests more neurites / glia, consistent with healthy white matter |
| ISOVF | Isotropic volume fraction | Extracellular water diffusion | More water outside cells suggests fewer neurites / glia, consistent with decreased health of white matter |

Sources for information in this table: (Cox et al., 2016; Elliott et al., 2018; Suzuki et al., 2017; Zhang et al., 2012)

Supplemental methods 8. Summary of IDPs included in analysis

We selected a subset of IDPs aiming to capture a range of brain measures while avoiding redundant measures. Specifically, our analysis included 354 IDPs: 4 represent total brain volumes from T1 MRI; 186 represent grey matter volume, area, and thickness in 62 cortical regions derived from T1 MRI using the Freesurfer Desikan Killiany parcellation (Smith et al., 2020); 36 represent regional subcortical volumes derived from T1 MRI using the Freesurfer ASEG parcellation (Fischl et al., 2002; Smith et al., 2020); and 14 IDPs are derived from T2 MRI and represent magnetic susceptibility in subcortical brain structures. We also included 108 IDPs derived from diffusion MRI representing four measures reflecting structural integrity in 27 white matter tracts. These IDPs are the diffusion tensor imaging (DTI) outputs fractional anisotropy (FA) and mean diffusivity (MD), and neurite orientation dispersion and density (NODDI) generated the measures intracellular volume fraction (ICVF) and isotropic volume fraction (ISOVF). We include 6 independent components derived from rfMRI in our analysis. The UK Biobank contains 1,695 rfMRI IDPs which represent the connectivity between a pair of brain regions. To simplify analysis, we performed independent component analysis (ICA) to reduce these to 6 independent components (ICs) representing broad patterns of connectivity through the brain using the procedure and weights provided by Elliot et al 2018 (Elliott et al., 2018).

| T1 | | T2 | | dMRI | | rfMRI | |
|---|---|---|---|---|---|---|---|
| Freesurfer ASEG[1] total volumes | 3 | Median T2* | 14 | Weighted mean FA | 27 | Independent components | 6 |
| Derived (total white matter) | 1 | | | Weighted mean MD | 27 | | |
| Freesurfer DKT[2] volume | 62 | | | Weighted mean ICVF | 27 | | |
| Freesurfer DKT[2] thickness | 62 | | | Weighted mean ISOVF | 27 | | |
| Freesurfer DKT[2] area | 62 | | | | | | |
| Freesurfer ASEG[1] (subcortical) | 36 | | | | | | |
| | | | | | | **Total IDPs** | **354** |

[1]Freesurfer ASEG is a parcellation dividing the subcortex into regions created using Freesurfer's automated segmentation and based on a brain atlas described by Fischl et al (Fischl et al., 2002)
[2]Freesurfer DKT divides the cortex into regions based on the Desikan Killiany brain atlas (Desikan et al., 2006; Fischl et al., 2004)

Supplemental methods 9. Map of Desikan Killiany parcellation with region names

Of the IDPs derived from T1 structural MRI, 186 IDPs represent area, thickness, and volume 62 in cortical regions defined by the Desikan Killiany atlas (Desikan et al., 2006; Destrieux et al., 2010; Fischl et al., 2004; Smith et al., 2020). The below map shows the spatial location of each of these regions. This map is from the ggseg3d package for R (Mowinckel, 2021; Mowinckel & Vidal-Piñeiro, 2020).

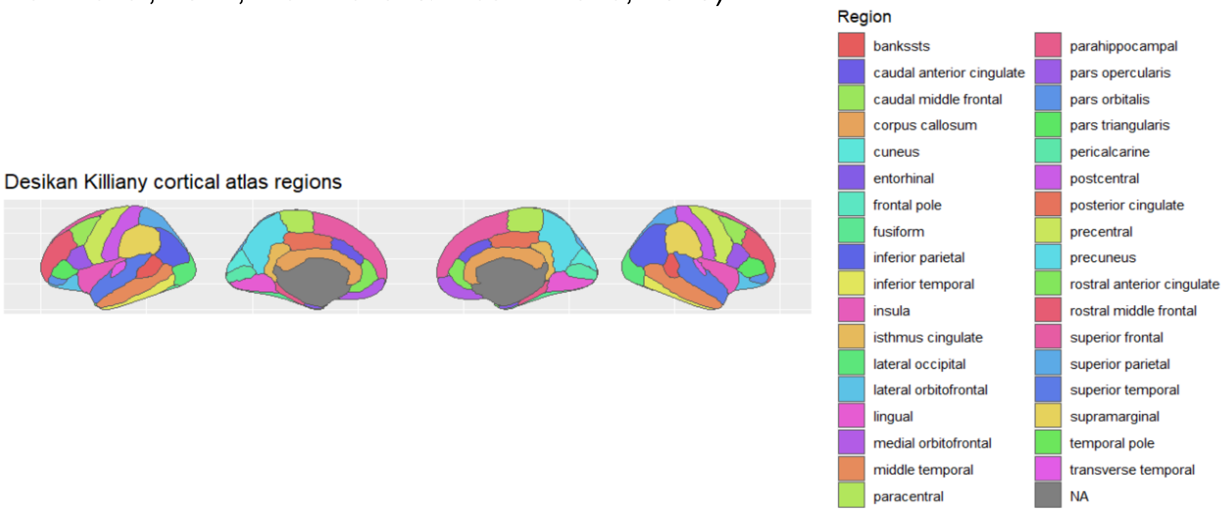

Supplemental methods 10. Map of Freesurfer ASEG subcortical parcellation with region names

UKB includes 36 IDPs representing volume of subcortical regions defined by the Freesurfer ASEG atlas (Fischl et al., 2002; Smith et al., 2020). Regions for which there are IDPs representing magnetic susceptibility from T2* can also be visualized on the same subcortical map, although they are not derived from Freesurfer ASEG. These maps are provided as a visualization tool to understand general spatial relationships and do not purport to be an accurate representation of neuroanatomy. This map is from the ggseg3d package for R (Mowinckel, 2021; Mowinckel & Vidal-Piñeiro, 2020) and we have added several additional regions for which the UKB has IDPs.

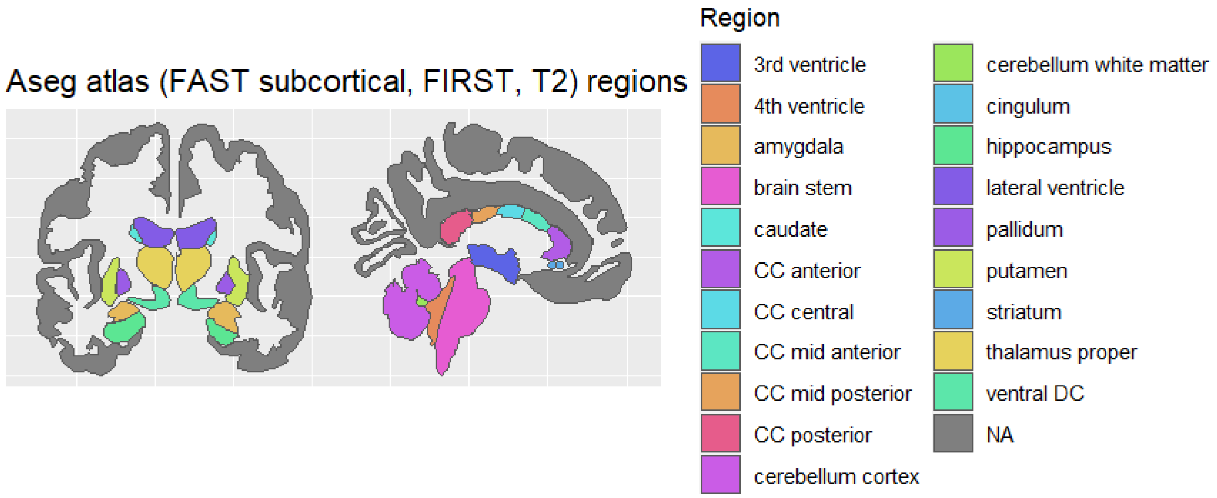

Supplemental methods 11. Map of 27 white matter tracts with names

UK Biobank dMRI IDPs are mapped onto 27 white matter tracts (3 medial and 13 bilateral). These are visualized using the atlas defined by Hua et al (Hua et al., 2008). We provide these maps as a visualization tool to understand general spatial relationships and do not purport them to be an accurate representation of neuroanatomy. This map is from the ggseg3d package for R (Mowinckel, 2021; Mowinckel & Vidal-Piñeiro, 2020) and we have added several additional regions for which the UKB has IDPs.

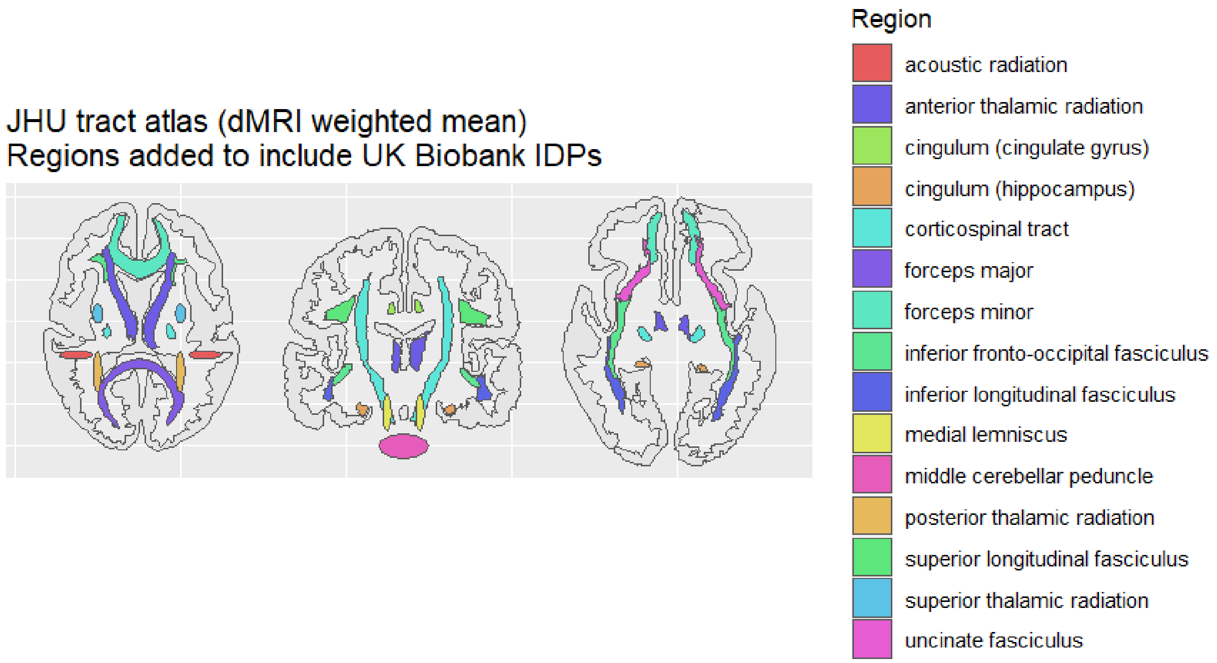

Supplemental methods 12. Flow through UKB alcohol touchscreen questions

In the UKB touchscreen survey, participants are first asked how frequently they consume alcohol (Data Field 1558). Participants who indicate consumption on a weekly basis are asked to estimate how much of several alcohol types they consume in the typical week. Those who indicate consumption less than weekly are asked to estimate how much of several alcohol types they consume in the typical month. Drinks per week is derived by summing Data Fields 1568, 1578, 1588, 1598, 1608, 5364 (weekly estimates by type of alcohol) and 4407, 4418, 4429, 4440, 4451, 4462 (monthly estimates by type of alcohol, divided by weeks in an average month). Participants who indicate that they never consume alcohol are asked if they formerly drank (Data Field 3731). This measure was z-score normalized in the linear regression model unless noted otherwise.

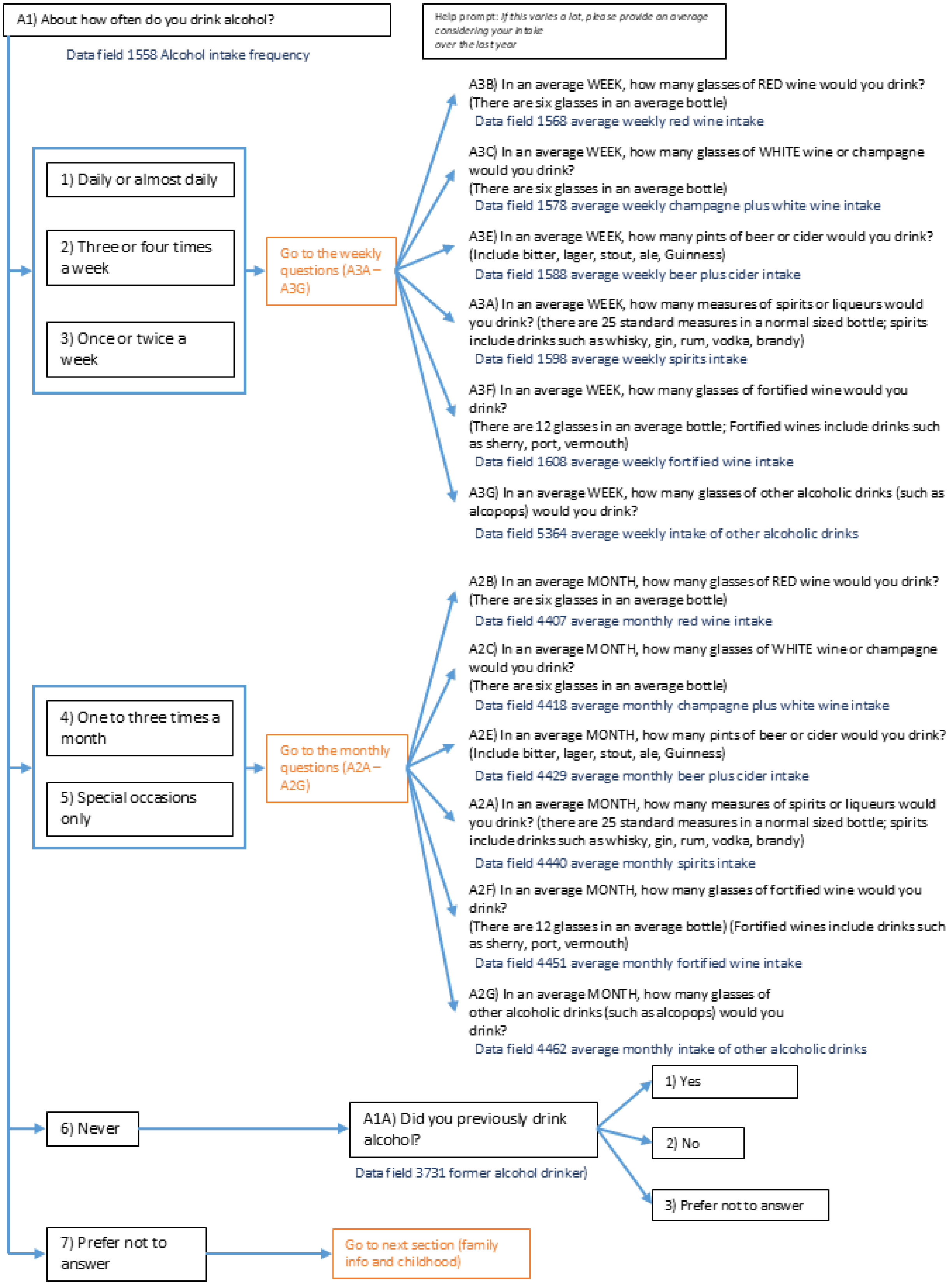
A1) About how often do you drink alcohol?
Data field 1558 Alcohol intake frequency
Help prompt: If this varies a lot, please provide an average considering your intake over the last year
1) Daily or almost daily
2) Three or four times a week
3) Once or twice a week
Go to the weekly questions (A3A – A3G)
A3B) In an average WEEK, how many glasses of RED wine would you drink? (There are six glasses in an average bottle)
Data field 1568 average weekly red wine intake
A3C) In an average WEEK, how many glasses of WHITE wine or champagne would you drink? (There are six glasses in an average bottle)
Data field 1578 average weekly champagne plus white wine intake
A3E) In an average WEEK, how many pints of beer or cider would you drink? (Include bitter, lager, stout, ale, Guinness)
Data field 1588 average weekly beer plus cider intake
A3A) In an average WEEK, how many measures of spirits or liqueurs would you drink? (there are 25 standard measures in a normal sized bottle; spirits include drinks such as whisky, gin, rum, vodka, brandy)
Data field 1598 average weekly spirits intake
A3F) In an average WEEK, how many glasses of fortified wine would you drink? (There are 12 glasses in an average bottle; Fortified wines include drinks such as sherry, port, vermouth)
Data field 1608 average weekly fortified wine intake
A3G) In an average WEEK, how many glasses of other alcoholic drinks (such as alcopops) would you drink?
Data field 5364 average weekly intake of other alcoholic drinks
4) One to three times a month
5) Special occasions only
Go to the monthly questions (A2A – A2G)
A2B) In an average MONTH, how many glasses of RED wine would you drink? (There are six glasses in an average bottle)
Data field 4407 average monthly red wine intake
A2C) In an average MONTH, how many glasses of WHITE wine or champagne would you drink? (There are six glasses in an average bottle)
Data field 4418 average monthly champagne plus white wine intake
A2E) In an average MONTH, how many pints of beer or cider would you drink? (Include bitter, lager, stout, ale, Guinness)
Data field 4429 average monthly beer plus cider intake
A2A) In an average MONTH, how many measures of spirits or liqueurs would you drink? (there are 25 standard measures in a normal sized bottle; spirits include drinks such as whisky, gin, rum, vodka, brandy)
Data field 4440 average monthly spirits intake
A2F) In an average MONTH, how many glasses of fortified wine would you drink? (There are 12 glasses in an average bottle) (Fortified wines include drinks such as sherry, port, vermouth)
Data field 4451 average monthly fortified wine intake
A2G) In an average MONTH, how many glasses of other alcoholic drinks (such as alcopops) would you drink?
Data field 4462 average monthly intake of other alcoholic drinks
6) Never
A1A) Did you previously drink alcohol?
Data field 3731 former alcohol drinker)
1) Yes
2) No
3) Prefer not to answer
7) Prefer not to answer
Go to next section (family info and childhood)

Supplemental methods 13. Flow through UKB smoking touchscreen questions

In the UKB touchscreen survey, participants are asked about current smoking (Data Field 1239). If they did not currently smoke daily, they were asked about past smoking (Data Field 1249). If participants indicated smoking on a daily basis currently or in the past, they were asked about smoking onset, offset, and packs per day to calculate pack years. Participants with less than daily smoking were asked if they had smoked over 100 cigarettes in their lifetime (Data Field 2644). Because participants with valid responses but no history of daily smoking were not presented with the questions to calculate pack years, we assigned 0 pack years to those who never smoked (less than 100 cigarettes in lifetime) and 1 pack year to those with occasional but never daily smoking. This measure was z-score normalized in the linear regression model unless noted otherwise.

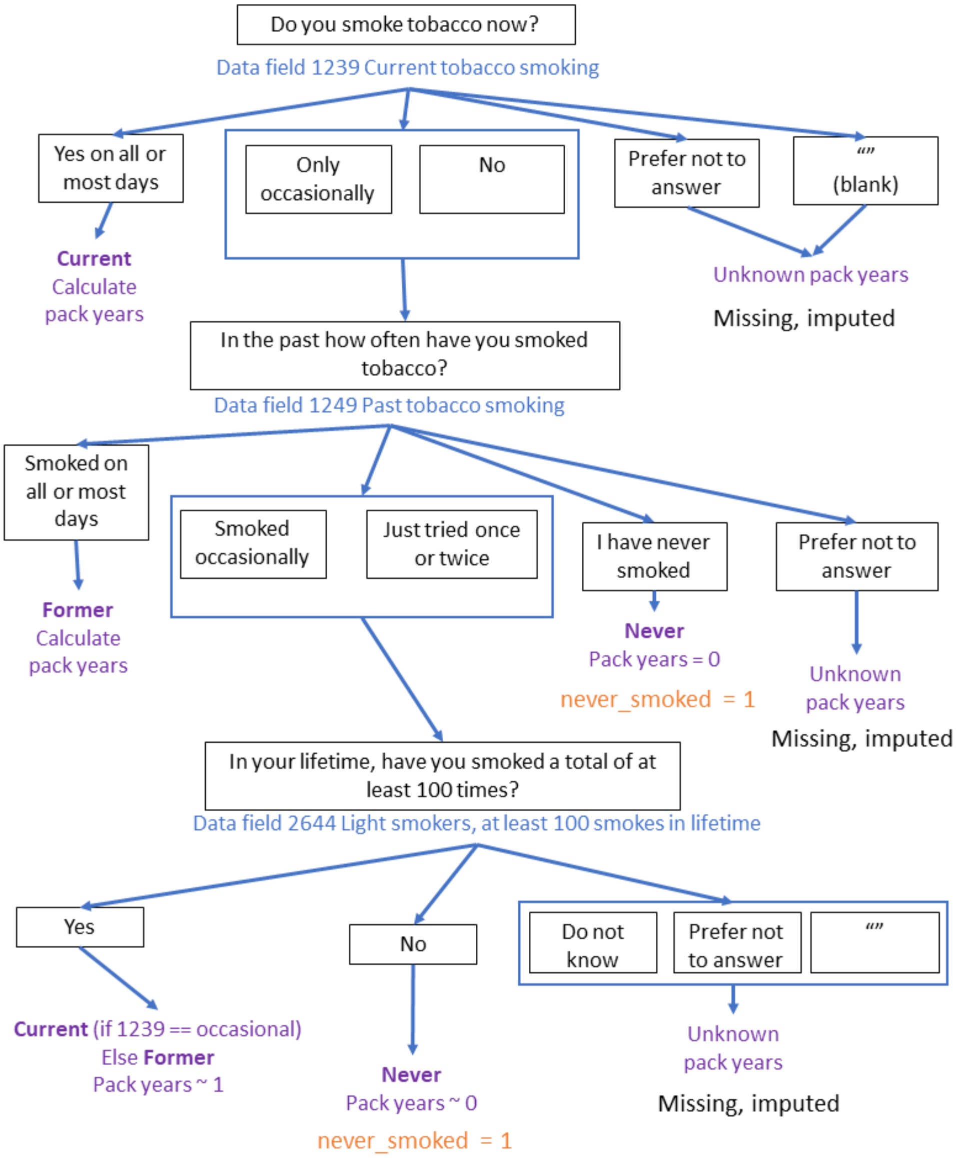

Supplemental methods 14. Main analysis regression model

Our goal was to include covariates relevant to brain health and substance use behavior while also maintaining a relatively parsimonious model without major concerns for collinearity between predictors. To select covariates for the model we referenced UKB neuroimaging literature (Alfaro-Almagro et al., 2021) which recommended the inclusion of head size, sex, age, imaging site, imaging date, and rfMRI derived motion. We included the health-related variables of body mass index (BMI), history of diabetes, systolic blood pressure (BP) and diastolic BP (Cole, 2020; Cox et al., 2019). We further included education (Davies et al., 2016; Zhou et al., 2021) and income (Dougherty et al., 2020; Shen et al., 2018; Weiss et al., 2024) to capture demographics. We determined that all these variables are associated with differences in total brain volume, contribute to model fit assessed with Akaike information criterion (AIC), and do not have variance inflation factor (VIF) > 5. Since we include participants of all races and ethnic backgrounds we also include the first 10 genetic principal components (PC).

IDP ~ drinks per week + pack years + drinks per week*pack years + sex + age + income + education + BMI + diabetes + systolic BP + diastolic BP + head size + imaging site + imaging date + rfMRI motion + PC1 + PC2 + PC3 + PC4 + PC5 + PC6 + PC7 + PC8 + PC9 + PC10

See supplemental methods 15 “Source and processing of model covariates other than alcohol and smoking” for details on the model covariates.

Supplemental methods 15. Source and processing of model covariates other than alcohol and smoking

This section describes the source and processing of the other covariates included in our regression model as controls. Sex, age, head size, imaging site, imaging date, and a variable representing how much the participant moved around in the scanner (rfMRI motion) are among the covariates recommended for analysis UKB imaging (Alfaro-Almagro et al., 2021). We additionally included the health-related covariates history of diabetes, body mass index (BMI), and systolic blood pressure (BP) and diastolic BP. Since we include participants of all races and ethnicities in our analysis we further include the first 10 genetic principle components.

Genetic sex, Data Field 22001
UKB offers two Data Fields for sex. We took genetic sex from Data Field 22001, sex determined from genetic analysis, as our measure of sex. Data Field 31 is sex taken from the central registry and updated by the participant during the baseline touchscreen survey, making it effectively self-reported sex. There were 21 participants in our imaging set for whom self-reported sex did not match genetic sex. We did not drop these participants from the sample but continued to classify them according to their genetic

sex. Sex was represented in the model as a binary variable (0 = female, 1 = male) (https://biobank.ndph.ox.ac.uk/showcase/search.cgi?wot=0&srch=22001&yfirst=2000&ylast=2024).

Age, Data Field 21003
Age of the participant at the neuroimaging appointment. This is a continuous numeric variable and is z-score normalized to a mean of 0 and standard deviation of 1 for the model (https://biobank.ndph.ox.ac.uk/showcase/field.cgi?id=21003).

Income, Data Field 738
Income is recorded in Data Field 738 “total household income before taxes” and is measured in pounds. Participants indicated which income band they fell into: less than 18,000, 18,000 to 30,999, 31,000 to 51,999, 52,000 to 100,000, and greater than 100,000 pounds. We converted this into a continuous numeric value by taking the lower bound of the income band. We performed the conversion on both the imaging visit and baseline values. Responses of “Do not know” and “Prefer not to answer” are treated as missing (https://biobank.ndph.ox.ac.uk/showcase/field.cgi?id=738).

Educational Attainment (Qualifications), Data Field 6138
We referenced Data Field 6138 “Which of the following qualifications do you have” to determine education level. We converted this into numeric years of education using the conversion system developed by Zhou et al and based on the International Standard Classification of Education (Zhou et al., 2021). Responses of “None of the above” and “Prefer not to answer” are treated as missing (https://biobank.ndph.ox.ac.uk/showcase/field.cgi?id=6138).

Body mass index (BMI), Data Field 21001
Participant height (Data Field 50) and weight (Data Field 21002) were collected at both the baseline and imaging visits used to calculate BMI (Data Field 21001). BMI is available for both instances. This is a continuous numeric variable and is z-score normalized to a mean of 0 and standard deviation of 1 for the model. (https://biobank.ndph.ox.ac.uk/showcase/field.cgi?id=21001)

History of diabetes, Data Field 2443
Participants were asked if they had ever been told by a doctor that they have diabetes (https://biobank.ndph.ox.ac.uk/showcase/field.cgi?id=2443). We took those who indicated “Yes” as having a history of diabetes. We encode this as a binary variable with 1 = history of diabetes. Responses of “Do not know” and “Prefer not to answer” were treated as missing values.

Systolic blood pressure (BP), Data Fields 4080 and 93
Data Field 4080 reflects systolic BP from a reading obtained with an automatic blood pressure cuff (https://biobank.ndph.ox.ac.uk/showcase/field.cgi?id=4080). When the automatic blood pressure cuff failed to get a reading the blood pressure was taken manually and recorded in field 93 (https://biobank.ndph.ox.ac.uk/showcase/field.cgi?id=93). If the automatic reading is

missing, we used values from the baseline instance, backfilling first with the measurement from the automatic blood pressure cuff reading and if that was missing, then the manual measurement. If all automatic and manual readings were missing, it was treated as a missing value. This is a continuous numeric variable and is z-score normalized to a mean of 0 and standard deviation of 1 for the model.

Diastolic blood pressure (BP), Data Fields 4079 and 94
Data Field 4079 reflects diastolic BP from a reading obtained with an automatic blood pressure cuff (https://biobank.ndph.ox.ac.uk/showcase/field.cgi?id=4079). When the automatic blood pressure cuff failed to get a reading the blood pressure was taken manually and recorded in field 94 (https://biobank.ndph.ox.ac.uk/showcase/field.cgi?id=94). If the automatic reading is missing, we used values from the baseline instance, backfilling first with the measurement from the automatic blood pressure cuff reading and if that was missing, then the manual measurement. If all automatic and manual readings were missing, it was treated as a missing value. This is a continuous numeric variable and is z-score normalized to a mean of 0 and standard deviation of 1 for the model.

Head size, Data Field 25000
While this variable is referred to as head size in descriptions of the model, it is volumetric scaling from T1 head imaging to volumetric space. This is a continuous numeric variable, and it is z-score normalized to a mean of 0 and standard deviation of 1 for the model (https://biobank.ndph.ox.ac.uk/showcase/field.cgi?id=25000).

Imaging site, Data Field 54
Out of all UKB data collection sites, imaging is conducted at 4. We represented the imaging site as a categorical variable with site 1 (Cheadle) as the reference (https://biobank.ndph.ox.ac.uk/showcase/field.cgi?id=54).

Imaging date, Data Field 53, instance 2
The date the participant came to the assessment center is recorded in Data Field 53. We reference instance 2 of this Data Field, which is the date the participant was imaged. We converted date of attending the imaging appointment into a numeric using an R function which transforms it into the number of days since 1 January 1970. This is a continuous numeric variable, and it is z-score normalized to a mean of 0 and standard deviation of 1 for the model (https://biobank.ndph.ox.ac.uk/showcase/field.cgi?id=53).

rfMRI motion, Data Field 25741
This variable represents head motion, averaged across space and time points (https://biobank.ndph.ox.ac.uk/showcase/field.cgi?id=25741).

Genetic principal components
The DNA samples collected at the baseline appointment were sent for genotyping using high quality standardized measures. The genotypes were processed and imputed to the Haplotype Reference Consortium in accordance with UK Biobank methods and made available. 40 genetic PCs were calculated for all participants and provided in Data Field

22009 “genetic principal components” (Bycroft et al., 2018). We included the first 10 of these in our regression model.

Supplemental methods 16. Backfilling and imputation of missing data

Participants missing the neuroimaging data (IDPs) or neuroimaging recommended covariates were dropped from our study cohort. We applied this missing data procedure to drinks per week, pack years, history of diabetes, body mass index (BMI), diastolic BP, systolic BP, education, and income.

Backfilling
Many Data Fields in the UKB have been collected twice, first at the baseline appointment and again at the imaging appointment. If available, we referenced the value from the imaging visit. The baseline data presented an opportunity to replace values missing from the imaging appointment instance with values collected during the baseline appointment “backfilling”. This greatly reduced the number of missing values which needed to be imputed.

Imputation
If a participant was missing a value from both the imaging and baseline visits, we imputed it using multiple imputation by chained equations (MICE) (van Buuren & Groothuis-Oudshoorn, 2011).

Supplemental methods 17. N of missing values before and after backfilling
“Backfilling” is described above in supplemental methods 16 “Backfilling and imputation of missing data”.

| Variable | Description | Derived from | N Missing Before backfilling | N Missing After* backfilling |
|---|---|---|---|---|
| week_drinks | Drinks per week | ** | 265 | 34 |
| pack_years | Pack years smoking | 20161 | 1319 | 400 |
| sex | Sex (genetic) | 22001 | 0 | 0 |
| age | Age at imaging appointment (years) | 21003 | 0 | 0 |
| income | Income (pounds) | 738 | 3,723 | 1,988 |
| BMI | Body mass index | 21001 | 1,295 | 0 |
| diabetes | Diabetes diagnosed by a doctor | 2443 | 350 | 4 |
| systolic_BP | Systolic blood pressure (mmHg) | 4080 (automatic), 93 (manual) | 4,695 | 2 |
| diastolic_BP | Diastolic blood pressure (mmHg) | 4079 (automatic), 94 (manual) | 4,695 | 2 |
| head_size | Volumetric scaling from T1 to standard space | 25000 | 0 | 0 |
| site | Imaging site | 54 | 0 | 0 |
| date | Imaging date (days since January 1 1970) | 53 | 0 | 0 |
| rfMRI_motion | Mean rfMRI motion | 25741 | 0 | 0 |

*Values that were missing after backfilling were imputed
**See supplemental methods 12 for derivation of alcohol measure from UKB data fields

Supplemental methods 18. Regression model covariates variance inflation factor (VIF)

In order to determine if collinearity between model covariates is a concern which may lead to instability of estimates we determined variance inflation factor (VIF) for all model covariates using the vif function from the cars library for r (https://www.rdocumentation.org/packages/car/versions/3.1-2/topics/vif). These are the results from the model fitted on total brain volume from Freesurfer ASEG, IDP 26514 (https://biobank.ndph.ox.ac.uk/showcase/field.cgi?id=26514). VIF less than 5 is generally considered acceptable and without major concerns for collinearity.

| Term | DF | VIF | Term | DF | VIF |
|---|---|---|---|---|---|
| week drinks | 1 | 1.19 | date | 1 | 1.56 |
| pack years | 1 | 1.19 | rfMRI motion | 1 | 1.68 |
| interaction | 1 | 1.18 | PC 1 | 1 | 1.08 |
| sex | 1 | 1.78 | PC 2 | 1 | 1.09 |
| age | 1 | 1.44 | PC 3 | 1 | 1.07 |
| income | 1 | 1.23 | PC 4 | 1 | 1.11 |
| education years | 1 | 1.11 | PC 5 | 1 | 1.06 |
| BMI | 1 | 1.67 | PC 6 | 1 | 1.02 |
| diabetes | 1 | 1.06 | PC 7 | 1 | 1.08 |
| systolic BP | 1 | 1.92 | PC 8 | 1 | 1.05 |
| diastolic BP | 1 | 1.78 | PC 9 | 1 | 1.04 |
| head size | 1 | 1.74 | PC 10 | 1 | 1.06 |
| site | 3 | 1.79 | | | |

Supplemental methods 19. Full demographics

| | | n | % |
|---|---|---|---|
| Sex[1] | Female | 19,220 | 53 |
| | Male | 17,089 | 47 |
| Age | <60 | 11,107 | 31 |
| | 60 - 69 | 15,455 | 43 |
| | 70+ | 9,747 | 27 |
| Race / ethnicity[2] | White | 35,151 | 97 |
| Income | <18,000 | 4,488 | 12 |
| | 18,000-30,999 | 9,958 | 27 |
| | 31,000-51,999 | 11,137 | 31 |
| | 52,000-100,000 | 8,139 | 22 |
| | >100,000 | 2,587 | 7 |
| Education | <10 years | 2,271 | 6 |
| | 10-16 years (CSEs or O levels and above) | 13,976 | 38 |
| | >16 years (college or university degree) | 20,062 | 55 |
| Alcohol use[3] | Never | 1,179 | 3 |
| | Current | 35,130 | 97 |
| Drinks per week | 0 (never) | 1,179 | 3 |
| | <=1 | 5,652 | 16 |
| | >1 - 7 | 15,175 | 42 |
| | >7 - 14 | 8,801 | 24 |
| | >14 - 21 | 2,913 | 8 |
| | >21 | 2,589 | 7 |
| Smoking[4] | Never | 22,818 | 63 |
| | Former | 12,255 | 34 |
| | Current | 1,236 | 3 |
| Pack years smoking | 0 (never) | 22,818 | 63 |
| | <=1 (infrequent) | 4,788 | 13 |
| | >1 - 10 | 2,976 | 8 |
| | >10 - 20 | 2,609 | 7 |
| | >20 - 40 | 2,353 | 6 |
| | >40 | 765 | 2 |
| BMI | <=18.5 (underweight) | 252 | 1 |
| | >18.5 - 24.9 (normal) | 14,257 | 39 |
| | >24.9 - 29.9 (overweight) | 15,214 | 42 |
| | >29.9 - 39.9 (obesity) | 6,218 | 17 |
| | >=40 (extreme obesity) | 368 | 1 |
| History of diabetes | % yes | 1,804 | 5 |
| Systolic BP | <=120 (normal) | 5,070 | 14 |
| | >120 - 129 (elevated) | 6,081 | 17 |
| | >129 - 139 (high, stage 1) | 7,340 | 20 |
| | >139 (high, stage 2) | 17,818 | 49 |
| Diastolic BP | <=80 (normal / elevated) | 20,425 | 56 |
| | >80 - 90 (high, stage 1) | 10,550 | 29 |
| | >90 (high, stage 2) | 5,334 | 15 |

[1]Sex is genetic sex. Sex from the National Health Service registry and corrected by participants during the baseline touchscreen survey is also available. 21 participants in the imaging cohort have genetic sex that does not match self-report.
[2]Ethnicity was self-reported during the touchscreen survey.
[3]Participants who formerly drank alcohol but indicate no current alcohol consumption are not included in the study population.
[4]Participants who indicated no or occasional smoking, and who indicated smoking less than 100 cigarettes in their lifetime are considered to have never smoked.

Supplemental methods 20. Relationship between drinks per week and pack years smoking

It has been reported that there may be substantial correlation between level of alcohol use and smoking, which could pose a threat to the validity of our model (Beard et al., 2017). To investigate this concern, we calculated the correlation between alcohol use as drinks per week and smoking as pack years (r = 0.18, p = 1.4*10^-262, n = 36309). This highly significant, low correlation supports the validity of findings from our model. When we stratify participants by drinks per week and calculate average pack years, we do see increased pack years in the higher alcohol consumption groups as would be expected.

Supplemental methods 21. Average pack years in groups defined by drinks per week

| Drinks per week | N | % | Pack years (mean) | Lower bound | Upper bound |
|---|---|---|---|---|---|
| 0 (never) | 1,179 | 3 | 2.63 | 2.04 | 3.22 |
| <=1 | 5,655 | 15 | 3.8 | 3.52 | 4.08 |
| (1,7] | 15,172 | 41 | 3.41 | 3.26 | 3.56 |
| (7,14] | 8,800 | 24 | 4.83 | 4.61 | 5.06 |
| (14,21] | 2,914 | 8 | 7.23 | 6.76 | 7.7 |
| >21 | 2,589 | 7 | 10.63 | 10 | 11.26 |

N out of 36,309 participants

Supplemental methods 22. Average drinks per week in groups defined by pack years

| Pack years | n | percent | Drinks per week (mean) | Lower bound | Upper bound |
|---|---|---|---|---|---|
| 0 (never*) | 22,813 | 62 | 6.43 | 6.34 | 6.52 |
| <=1** | 4,792 | 13 | 9.14 | 8.9 | 9.38 |
| (1,10] | 2,966 | 8 | 9.8 | 9.48 | 10.12 |
| (10,20] | 2,612 | 7 | 10.57 | 10.18 | 10.95 |
| (20,40] | 2,356 | 6 | 11.49 | 11.05 | 11.93 |
| >40 | 770 | 2 | 12.62 | 11.66 | 13.57 |

N out of 36,309 participants
*Participants who indicated smoking less than 100 cigarettes in their lifetime in Data Field 2644 were assigned 0 pack years and considered to have never smoked.
**Participants who indicated no history of daily smoking but more than 100 cigarettes in their lifetime were not asked about pack years in the touchscreen survey. We assigned these participants 1 pack year to represent their occasional, but less than daily, smoking history.

Supplemental methods 23. References

Alfaro-Almagro, F., McCarthy, P., Afyouni, S., Andersson, J. L. R., Bastiani, M., Miller, K. L., Nichols, T. E., & Smith, S. M. (2021). Confound modelling in UK Biobank brain imaging. *NeuroImage*, *224*(2021), 117002. https://doi.org/10.1016/j.neuroimage.2020.117002

Beard, E., West, R., Michie, S., & Brown, J. (2017). Association between smoking and alcohol-related behaviours: A time–series analysis of population trends in England. *Addiction (Abingdon, England)*, *112*(10), 1832–1841. https://doi.org/10.1111/add.13887

Bycroft, C., Freeman, C., Petkova, D., Band, G., Elliott, L. T., Sharp, K., Motyer, A., Vukcevic, D., Delaneau, O., O'Connell, J., Cortes, A., Welsh, S., Young, A., Effingham, M., McVean, G., Leslie, S., Allen, N., Donnelly, P., & Marchini, J. (2018). The UK Biobank resource with deep phenotyping and genomic data. *Nature*, *562*(7726), 203–209. https://doi.org/10.1038/s41586-018-0579-z

Cole, J. H. (2020). Multimodality neuroimaging brain-age in UK biobank: Relationship to biomedical, lifestyle, and cognitive factors. *Neurobiology of Aging*, *92*, 34–42. https://doi.org/10.1016/j.neurobiolaging.2020.03.014

Cox, S. R., Lyall, D. M., Ritchie, S. J., Bastin, M. E., Harris, M. A., Buchanan, C. R., Fawns-Ritchie, C., Barbu, M. C., de Nooij, L., Reus, L. M., Alloza, C., Shen, X., Neilson, E., Alderson, H. L., Hunter, S., Liewald, D. C., Whalley, H. C., McIntosh, A. M., Lawrie, S. M., … Deary, I. J. (2019). Associations between vascular risk factors and brain MRI indices in UK Biobank. *European Heart Journal*, *40*(28), 2290–2300. https://doi.org/10.1093/eurheartj/ehz100

Cox, S. R., Ritchie, S. J., Tucker-Drob, E. M., Liewald, D. C., Hagenaars, S. P., Davies, G., Wardlaw, J. M., Gale, C. R., Bastin, M. E., & Deary, I. J. (2016). Ageing and brain white matter structure in 3,513 UK Biobank participants. *Nature Communications*, *7*(13629). https://doi.org/10.1038/ncomms13629

Davies, G., Marioni, R. E., Liewald, D. C., Hill, W. D., Hagenaars, S. P., Harris, S. E., Ritchie, S. J., Luciano, M., Fawns-Ritchie, C., Lyall, D., Cullen, B., Cox, S. R., Hayward, C., Porteous, D. J., Evans, J., McIntosh, A. M., Gallacher, J., Craddock, N., Pell, J. P., … Deary, I. J. (2016). Genome-wide association study of cognitive functions and educational attainment in UK Biobank (N=112 151). *Molecular Psychiatry*, *21*(6), 758–767. https://doi.org/10.1038/mp.2016.45

Desikan, R. S., Ségonne, F., Fischl, B., Quinn, B. T., Dickerson, B. C., Blacker, D., Buckner, R. L., Dale, A. M., Maguire, R. P., Hyman, B. T., Albert, M. S., & Killiany, R. J. (2006). An automated labeling system for subdividing the human cerebral cortex on MRI scans into gyral based regions of interest. *NeuroImage*, *31*(3), 968–980. https://doi.org/10.1016/j.neuroimage.2006.01.021

Destrieux, C., Fischl, B., Dale, A., & Halgren, E. (2010). Automatic parcellation of human cortical gyri and sulci using standard anatomical nomenclature. *NeuroImage*, *53*(1), 1–15. https://doi.org/10.1016/j.neuroimage.2010.06.010

Dougherty, R. J., Moonen, J., Yaffe, K., Sidney, S., Davatzikos, C., Habes, M., & Launer, L. J. (2020). Smoking mediates the relationship between SES and brain volume: The CARDIA study. *PloS One*, *15*(9), e0239548. https://doi.org/10.1371/journal.pone.0239548

Elliott, L. T., Sharp, K., Alfaro-Almagro, F., Shi, S., Miller, K. L., Douaud, G., Marchini, J., & Smith, S. M. (2018). Genome-wide association studies of brain imaging phenotypes in UK Biobank. *Nature*, *562*(7726), 210–216. https://doi.org/10.1038/s41586-018-0571-7

Fischl, B., Salat, D. H., Busa, E., Albert, M., Dieterich, M., Haselgrove, C., van der Kouwe, A., Killiany, R., Kennedy, D., Klaveness, S., Montillo, A., Makris, N., Rosen, B., & Dale, A. M. (2002). Whole Brain Segmentation: Automated Labeling of Neuroanatomical Structures in the Human Brain. *Neuron*, *33*(3), 341–355. https://doi.org/10.1016/S0896-6273(02)00569-X

Fischl, B., Van Der Kouwe, A., Destrieux, C., Halgren, E., Ségonne, F., Salat, D. H., Busa, E., Seidman, L. J., Goldstein, J., Kennedy, D., Caviness, V., Makris, N., Rosen, B., & Dale, A. M. (2004). Automatically Parcellating the Human Cerebral Cortex. *Cerebral Cortex*, *14*(1), 11–22. https://doi.org/10.1093/cercor/bhg087

Gray, J. C., Thompson, M., Bachman, C., Owens, M. M., Murphy, M., & Palmer, R. (2020). Associations of cigarette smoking with gray and white matter in the UK Biobank. *Neuropsychopharmacology*, *45*(7), 1215–1222. https://doi.org/10.1038/s41386-020-0630-2

Hanscombe, K. B., Coleman, J. R. I., Traylor, M., & Lewis, C. M. (2019). ukbtools: An R package to manage and query UK Biobank data. *PLoS ONE*, *14*(5), e0214311. https://doi.org/10.1371/journal.pone.0214311

Hua, K., Zhang, J., Wakana, S., Jiang, H., Li, X., Reich, D. S., Calabresi, P. A., Pekar, J. J., van Zijl, P. C. M., & Mori, S. (2008). Tract Probability Maps in Stereotaxic

Spaces: Analyses of White Matter Anatomy and Tract-Specific Quantification. *NeuroImage*, *39*(1), 336–347. https://doi.org/10.1016/j.neuroimage.2007.07.053

Mowinckel, A. M. (2021). *Plotting Tool for Brain Atlases*. https://doi.org/10.1177/2515245920928009>.License

Mowinckel, A. M., & Vidal-Piñeiro, D. (2020). Visualization of Brain Statistics With R Packages ggseg and ggseg3d. *Advances in Methods and Practices in Psychological Science*, *3*(4), 466–483. https://doi.org/10.1177/2515245920928009

Shen, X., Cox, S. R., Adams, M. J., Howard, D. M., Lawrie, S. M., Ritchie, S. J., Bastin, M. E., Deary, I. J., McIntosh, A. M., & Whalley, H. C. (2018). Resting-State Connectivity and Its Association With Cognitive Performance, Educational Attainment, and Household Income in the UK Biobank. *Biological Psychiatry: Cognitive Neuroscience and Neuroimaging*, *3*(10), 878–886. https://doi.org/10.1016/j.bpsc.2018.06.007

Smith, S. M., Alfaro-almagro, F., & Miller, K. L. (2020). *UK Biobank Brain Imaging Documentation*. https://www.ukbiobank.ac.uk/

Suzuki, H., Gao, H., Bai, W., Evangelou, E., Glocker, B., O'Regan, D. P., Elliott, P., & Matthews, P. M. (2017). Abnormal brain white matter microstructure is associated with both pre-hypertension and hypertension. *PLoS ONE*, *12*(11), e0187600. https://doi.org/10.1371/journal.pone.0187600

van Buuren, S., & Groothuis-Oudshoorn, K. (2011). mice: Multivariate Imputation by Chained Equations in R. *Journal of Statistical Software*, *45*, 1–67. https://doi.org/10.18637/jss.v045.i03

Weiss, J., Beydoun, M. A., Beydoun, H. A., Georgescu, M. F., Hu, Y.-H., Noren Hooten, N., Banerjee, S., Launer, L. J., Evans, M. K., & Zonderman, A. B. (2024). Pathways explaining racial/ethnic and socio-economic disparities in brain white matter integrity outcomes in the UK Biobank study. *SSM - Population Health*, *26*, 101655. https://doi.org/10.1016/j.ssmph.2024.101655

Zhang, H., Schneider, T., Wheeler-Kingshott, C. A., & Alexander, D. C. (2012). NODDI: Practical in vivo neurite orientation dispersion and density imaging of the human brain. *NeuroImage*, *61*(4), 1000–1016. https://doi.org/10.1016/j.neuroimage.2012.03.072

Zhou, T., Sun, D., Li, X., Ma, H., Heianza, Y., & Qi, L. (2021). Educational attainment and drinking behaviors: Mendelian randomization study in UK Biobank. *Molecular Psychiatry*, *26*(8), 4355–4366. https://doi.org/10.1038/s41380-019-0596-9
